# Epidemiological model with anomalous kinetics - Early Stages of the Covid-19 pandemic

**DOI:** 10.1101/2020.06.24.20139287

**Authors:** Ugur Tirnakli, Constantino Tsallis

## Abstract

We generalize the phenomenological, law of mass action-like, SIR and SEIR epidemiological models to situations with anomalous kinetics. Specifically, the contagion and removal terms, normally linear in the fraction *I* of infecteds, are taken to depend on 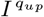 and 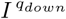, respectively. These dependencies can be understood as highly reduced effective descriptions of contagion via anomalous diffusion of susceptibles and infecteds in fractal geometries, and removal (i.e., recovery or death) via complex mechanisms leading to slowly decaying removal-time distributions. We obtain rather convincing fits to time series for both active cases and mortality with the same values of (*q*_*up*_, *q*_*down*_) for a given country, suggesting that such aspects may in fact be present in the early evolution of the Covid-19 pandemics. We also obtain approximate values for the effective population *N*_*eff*_, which turns out to be a small percentage of the entire population *N* for each country.

## GENERALIZED MODELS

### *q*-SIR Model

The SIR set of equations is (see [1] for instance) as follows:

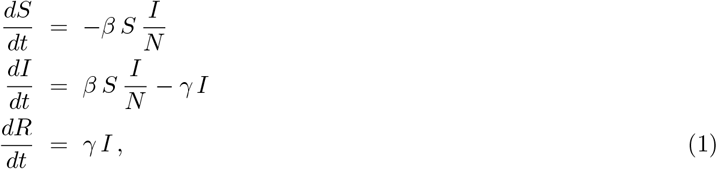

with *β* > 0, *γ* > 0, and *S* + *I* + *R* = *N* = *constant, N* is the total population, *S* ≡ *susceptible, I* ≡ *infected, R* ≡ *removed* (*removed* means either recovered or dead). Now let us *q*-generalize this model as follows:

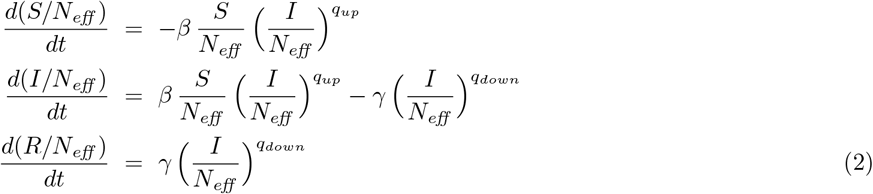

with *q*_*up*_ ≤ 1 and *q*_*down*_ ≥ 1, where the bilinear term is generalized into a non bilinear one and the *effective* population *N*_*eff*_ = *ρN* with *ρ* ≤ 1. These equations generically have a single peak for *I*(*t*). In all cases we have *S*(*t*) + *I*(*t*) + *R*(*t*) = *N*_*eff*_; moreover, 0 ≤ *S*(*t*)*/N*_*eff*_, *I*(*t*)*/N*_*eff*_, *R*(*t*)*/N*_*eff*_ ≤ 1. Consistently, in the set of equations 2, it is enough to retain the first two. Let us qualitatively compare the SIR and qSIR models given by Eqs. (1) and (2) respectively by focusing on the *β* term, i.e., let us compare *β*_*SIR*_ [*S*(*t*)*/N*] [*I*(*t*)*/N*] with 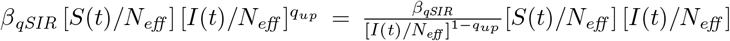; we remind that *ρ* = 1 yields *N*_*eff*_ = *N*. It follows that roughly 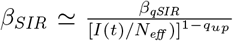. Since, before the peak, *I*(*t*) steadily *increases* with time, a fixed value for (1 − *q*_*up*_) > 0 acts qualitatively as a phenomenological time-dependent *β*_*SIR*_(*t*) which *decreases* with time. These tendencies are similarly realistic since they both reflect, each in its own manner, the generic action of pandemic authorities to isolate people in order to decrease the contagion represented by the *β* term in both models.

The particular limit *R*(*t*) ≡ 0 (hence *S*(*t*) + *I*(*t*) = *N*_*eff*_) in Eqs. (2) yields 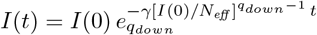 if *β* = 0, and 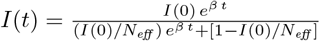 if (*γ, q*_*up*_) = (0, 1). For generic *q*_*up*_ < 1 and *R*(*t*) ≡ 0, Eqs. (2) yield

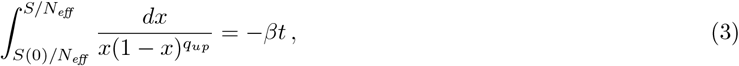

hence

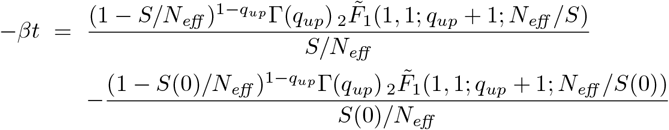

where Γ is the Gamma function and 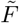 is the regularized hypergeometric function. As an illustration let us consider *q*_*up*_ = 1*/*2. It follows ln 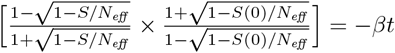, hence

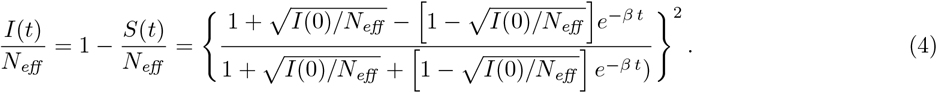

Before the peak, *I*(*t*) increases nearly exponentially if *q*_*up*_ = 1 and is roughly characterized by 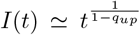 if *q*_*up*_ < 1. After the peak, *I*(*t*) decreases exponentially if *q*_*down*_ = 1 and is roughly characterized by 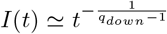 if *q*_*down*_ > 1. These various aspects are illustrated in Fig. 1. An important remark is necessary at this point. The possibility for non bilinear coupling 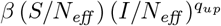 between subpopulations seems quite natural since non-homogeneous mixing involves complex dynamics and networks for the susceptible and infected people, as well as for the infecting agent of the disease. But why would it be necessary to also allow, at the present phenomenological level, a nonlinear behavior for the one-subpopulation term *γ* 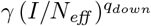 itself? The answer might be found in nontrivial (multi)fractal-path-like relaxation mechanisms such as the one that is known to happen in re-associations in folded proteins [16, 17].

**Figure 1:**
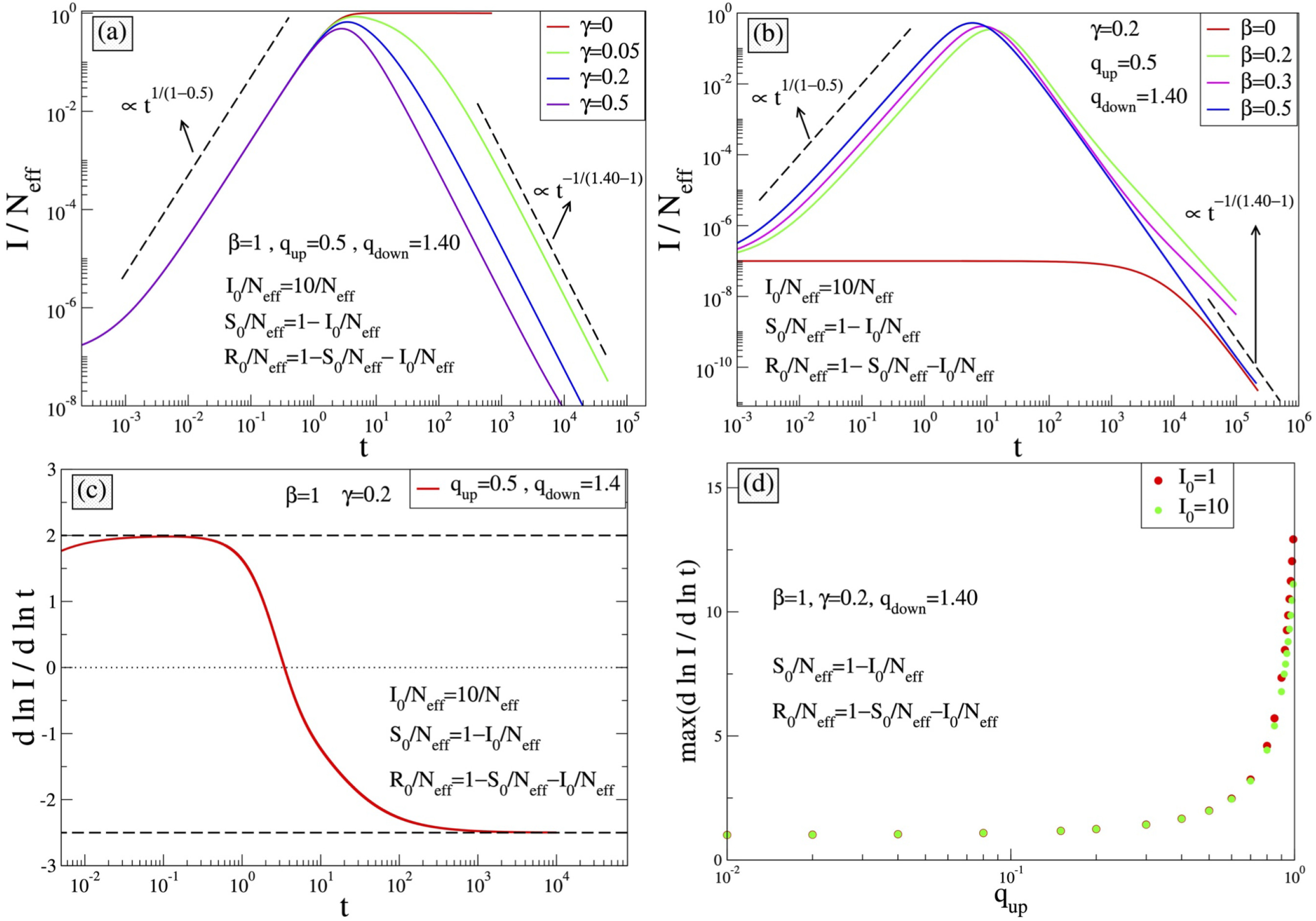
Time evolution of *I*(*t*) for the *q*-SIR equations with *N*_*eff*_ = 10^8^; we consider *t* to run virtually from zero to infinity (not necessarily during only the typical range of real epidemics, say 1000 days). (a) Fixed *β* and various values of *γ*. For *γ* = 0 and ∀*β, I*(*t*)*/N*_*eff*_ precisely recovers the expression given in Eq. (4). The slope at the first *inflexion point* for increasing time is given by *max*[*d* ln *I/d* ln *t*] = 1*/*(1 − *q*_*up*_), ∀ *γ*. (b) Fixed *γ* and various values of *β*. For *β* = 0, the *q*_*down*_-exponential function is precisely recovered. (c) Time dependence of the slope of [*d* ln(*I/N*_*eff*_)*/d* ln *t*], which appears to be bounded between 1*/*(1 − *q*_*up*_) (= 2 in this example) and −1*/*(*q*_*down*_ − 1) (= −2.5 in this example) (dashed lines). The dotted line corresponds to the position of the peak of *I*(*t*), and the maximum (minimum) corresponds to the left (right) inflexion point. (d) The maximal slope of *I*(*t*)*/N*_*eff*_ as a function of *q*_*up*_. The high value at *q*_*up*_ = 1 reflects the divergence expected in the limit *q*_*up*_ = *q*_*down*_ = 1. Indeed, in this limit, the present *q*-SIR model recovers the standard SIR model, which increases exponentially (and not as a power-law) towards the corresponding peak. Notice also that this slope decreases when *I*_0_*/N*_*eff*_ increases.

We have checked that the *q*-SIR model provides functions *I*(*t*) which are numerically close but different from the quite performing ansatz in [5], namely 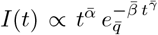, whose behavior before and after the peak are respectively 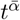 and 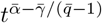. Still, both approaches have power-law behaviors before and after the peak. We also checked 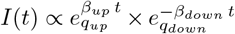, and the results are once again numerically close but nevertheless different from the ansatz in [5].

### *q*-SEIR Model

The *q*-SIR model is not capable (for any choice of its parameters) of correctly fitting the epidemiologically crucial function *I*(*t*) for the COVID-19 available data for various countries. Since this generalization of the simplest model does not provide a useful tool for COVID-19 data, we addressed a more sophisticated one, namely a four-compartment model known as SEIR. Therefore, we next *q*-generalize the SEIR model with no vital dynamics (no births, no deaths), which is given by

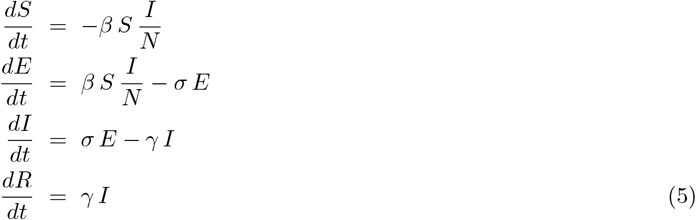

with *S* + *E* + *I* + *R* = *N*, where *E* stands for *exposed*. We can generalize it as follows:

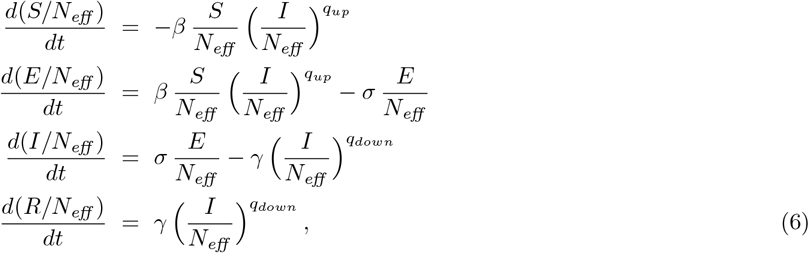

where once again we have generalized the bilinear couplings between subpopulations into non bilinear ones, and the linear *γ I* term into a nonlinear one. Its particular instance *E*(*t*) ≡ 0 (hence *S*(*t*) + *I*(*t*) + *R*(*t*) = *N*_*eff*_) precisely recovers the *q*-SIR model, as defined here above. Notice that the *cumulative function C*(*t*) of *I*(*t*) is given by 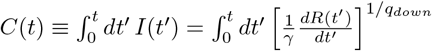, which differs from the expression *C*(*t*) = *R*(*t*) currently used in the SEIR model. It is of course possible to further generalize the above *q*-SEIR set of four equations by allowing in the right hand 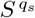 (with *q*_*s*_ ≠ 1) instead of *S*, and 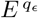 (with *q*_*ϵ*_ ≠ 1) instead of *E*, but no need has emerged to increase the number of free parameters of the model, since the allowance for *q*_*up*_ < 1 and for *q*_*down*_ > 1 appears to be enough for satisfactorily reproducing all the relevant features of the COVID-19 available data. Indeed, the variable which is epidemiologically crucial for avoiding a medical-hospital collapse in a given region is *I*(*t*), and this time dependence generically appears to be very satisfactorily described by just allowing the possibility for *q*_*up*_ ≠ 1 and/or *q*_*down*_ ≠1. Notice that 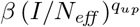 is a *convex* function of (*I/N*_*eff*_) for 0 < *q*_*up*_ < 1, and 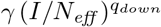 is a *concave* function of (*I/N*_*eff*_) for *q*_*down*_ > 1. These tendencies, illustrated in Fig. 2, as well as the numerical values for the various coefficients of the model, are in agreement with the available medical/epidemiological evidence [21–25]. Notice also that, through *τ* ≡ *γt* and 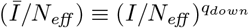, we can eliminate, without loss of generality, two fitting parameters (e.g., *γ* and *q*_*down*_) within the set of equations (6). Finally notice that, if we consider *β* = *σ* = 0, we precisely recover Eq. (**??**) and its analytical solution in Eq. (**??**). Therefore, even if the general analytical solution of the set of equations (6) is not available (due to mathematical intractability), the initials conditions naturally renormalize (in a nontrivial manner) the coefficients (*β, σ, γ*) of the model, and the effective population *N*_*eff*_ becomes a fitting parameter of the model. These renormalizations disappear of course if *q*_*up*_ = *q*_*down*_ = 1, i.e., for the standard SEIR model; concomitantly *N*_*eff*_ (= *ρN*, with *ρ* ≤ 1) ceases being a fitting parameter and can be directly taken from the actual population *N* of the particular region under focus. For other mathematical aspects of nonlinear models such as the present one, the reader may refer to [26].

**Figure 2:**
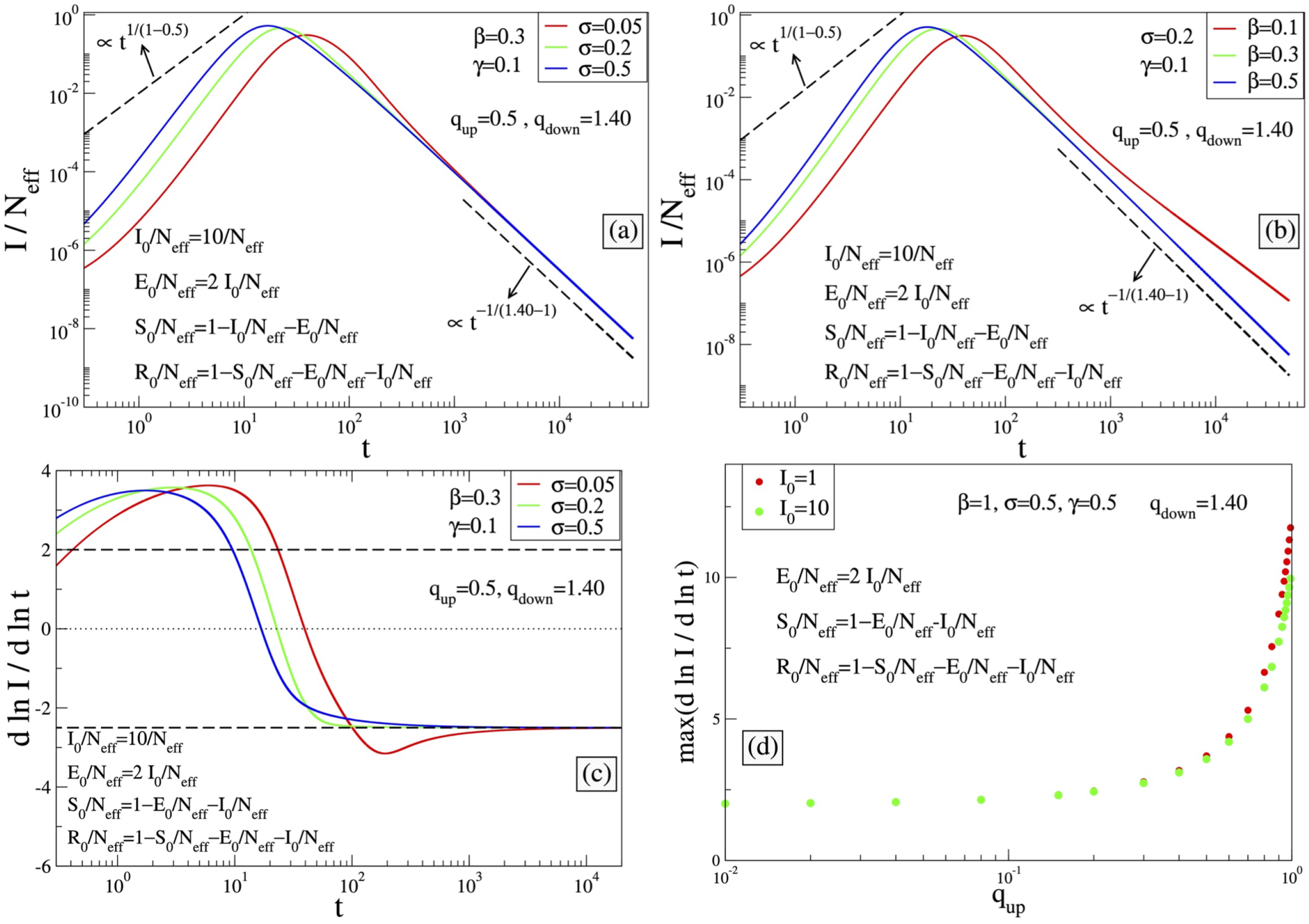
Time evolution of *I*(*t*) for the *q*-SEIR equations with *N*_*eff*_ = 10^8^; we consider *t* to run virtually from zero to infinity (not necessarily during only the typical range of real epidemics, say 1000 days). (a) Fixed (*β, γ*) and various values of *σ*. (b) Fixed (*σ, γ*) and various values of *β*. (c) Time dependence of the slope of [*d* ln *I/d* ln *t*], which, in contrast with the *q*-SIR equations, is *not* bounded between 1*/*(1 − *q*_*up*_) > 0 and − 1*/*(*q*_*down*_ − 1) < 0 (dashed lines). The dotted line corresponds to the position of the peak of *I*(*t*), and the maximum (minimum) corresponds to left (right) inflexion points. (d) The maximal slope of *I*(*t*)*/N*_*eff*_ as a function of *q*_*up*_. The high value at *q*_*up*_ = 1 reflects the divergence expected in the limit *q*_*up*_ = *q*_*down*_ = 1. Indeed, in this limit, the present *q*-SEIR model recovers the standard SEIR model, which increases exponentially (and not as a power-law) towards the corresponding peak. Notice also that this slope decreases when *I*_0_*/N*_*eff*_ increases.

## APPLICATION OF q-SEIR MODEL TO COVID-19 PANDEMICS

In Figs. 3 and 4 we have illustrations of this model for realistic COVID-19 cases. We identify the present variable *I* with the number of *active cases*[50], as regularly updated online [27]. We verify that the description provided by the *q*-SEIR model for non-homogeneous epidemiological mixing is indeed quite satisfactory for the early stages of the pandemics (before an unpredictable but possible second wave). Let us also mention that we have not followed here a road looking for the minimal number of free parameters, but rather a road where various realistic elements are taken into account, even if at the fitting-parameter level some of them might be redundant. Any further model yielding a deeper, or even first-principle, expression of exponents such as *q*_*up*_ and *q*_*down*_ in terms of microscopic/mesoscopic mechanisms is very welcome. This is by no means a trivial enterprise but, if successfully implemented, this would probably follow a road analogous to anomalous diffusion, namely from Fourier’s heat equation through Muskat’s *Porous Medium Equation* [28] to Plastino and Plastino nonlinear Fokker-Planck equation [29], which in turn implied the scaling law 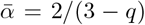 [30] (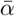being defined through the scaling between *x*^2^ and 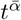, and *q* being the index value of the *q*-Gaussian solution for the nonlinear Fokker-Planck equation). This scaling law recovers, for *q* = 1, the Brownian motion scaling ⟨*x*^2^⟩ ∝ *t*, and was impressively validated within 2% error in granular matter [31]. This phenomenological line was later legitimated on the basis of microscopic overdamped mechanisms in at least a wide class of systems, namely providing *q* = 0 for the motion of vortices in type-II superconductors [32], later extended to *D*-dimensional 1*/r*^*λ*^ short-range repulsive interactions (*λ/D* ≥ 1), leading to *q* = 1 − *λ/D* [33]). These approaches were shown to satisfy the zeroth law of thermodynamics, an *H*-theorem, and Carnot’s cycle efficiency, with microscopically established analytical equations of state [34–37]. An attempt to follow along similar lines for the present *q*-SEIR model would surely be a very interesting challenge. Summarizing, we have *q*-generalized, through Eqs. (6), the SEIR epidemiological model. By solving this set of deterministic equations given the initial conditions and its parameters, we obtain [*S*(*t*), *E*(*t*), *I*(*t*)), *R*(*t*)], as well as the cumulative function *C*(*t*). We have focused on *I*(*t*) because the hardest quantities for satisfactorily fitting are the number of active cases and that of deaths, and also because those are the most crucial quantities for making correct sanitary and epidemiological decisions.

**Figure 3:**
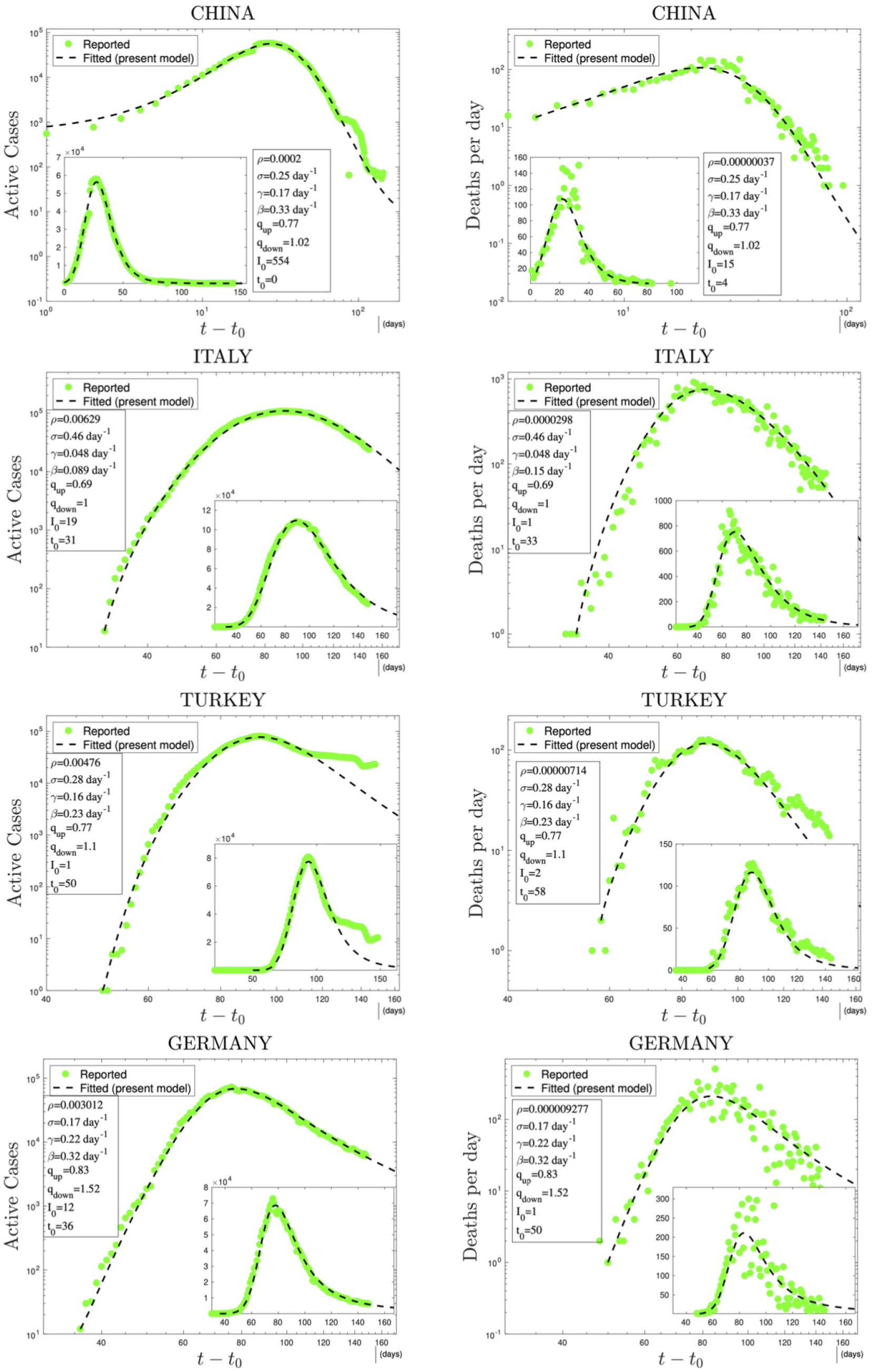
Time evolution of available data for COVID-19 numbers of active cases (probably under-reported in most cases) and deaths per day [27] and their (linear scale) Least Squares Method fittings with *N*_*eff*_ *I*(*t*) from the *q*-SEIR model; *ρ* = *N*_*eff*_ */N* where *N* is the population of the country. Notice that (i) By convention, *t*_0_ = 0 for China; (ii) Parameters such as *ρN* are particularly relevant for sanitary-epidemiological decisions, and, as it is natural, *ρ*(*deaths*) ≪ *ρ*(*active*) for any given country; (iii) For any given country, the values of (*q*_*up*_, *q*_*down*_) are the same for both curves of active cases and of deaths; (iv) The dates of the peaks of the active cases and deaths per day do not necessarily coincide; (v) The values that emerge for *β/γ* (*reproduction number* or *growth rate*), 1*/β* (*exposition time*), 1*/γ* (*recovering time*), and 1*/σ* (*incubation time*) are consistent with those currently indicated in the literature [21–25]; (vi) We considered all the data reported until June 13^*th*^, excluding some very initial transients or sudden anomalous discrepancies (e.g., in China, Turkey and Brazil).

**Figure 4:**
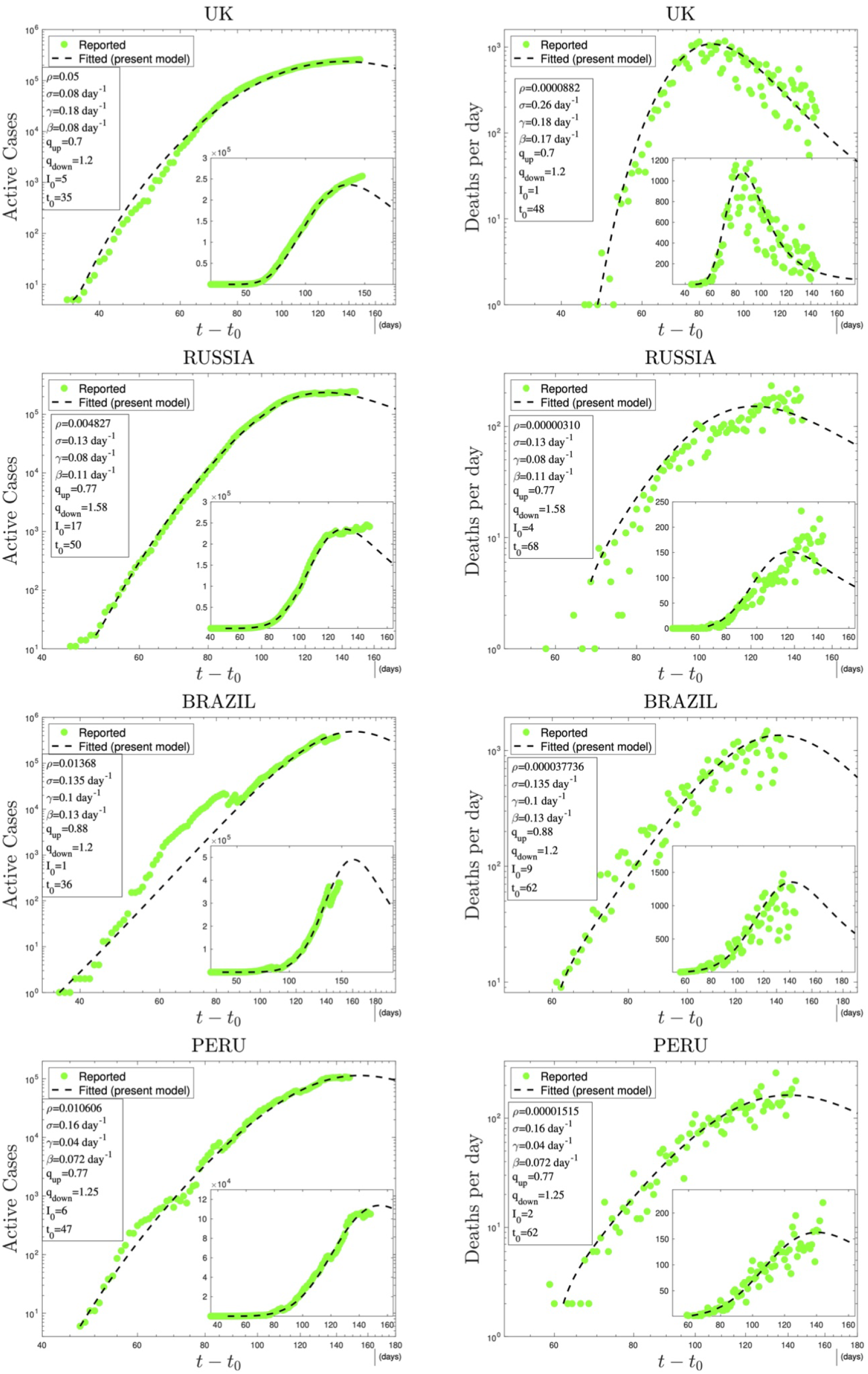
Continuation of figure.3

On a general perspective, let us stress that the law of mass action, the Arrhenius relaxation law, and the Kramers mechanism [38] of escape over a barrier through normal diffusion constitute pillars of contemporary chemistry. They are consistent with Boltzmann-Gibbs (BG) statistical mechanics and constitute some of its important successes. However, they need to be modified when the system exhibits complexity due to hierarchical space and/or time structures. It is along this line that a generalization has been proposed based on nonadditive entropies [20], characterized by the index *q* (*q* = 1 recovers the BG frame): see for instance [16, 39–43]. It is along these same lines that lies the present *q*-generalization of the standard SEIR model.

At the level of the numerical performance of the present *q*-SEIR model for the COVID-19 pandemic, it advantageously compares with models including time-dependent coefficients [44–47]. For instance, the SEIQRDP model [44, 45, 47] includes seven equations with several coefficients, two of them phenomenologically being time-dependent. It does fit rather well the COVID-19 reported data until a given date. However, the *q*-SEIR, which includes four (instead of seven) equations with several coefficients, all of them being fixed in time, fits definitively better the same data for all the countries that we have checked: see illustrations in Fig. 5.

**Figure 5:**
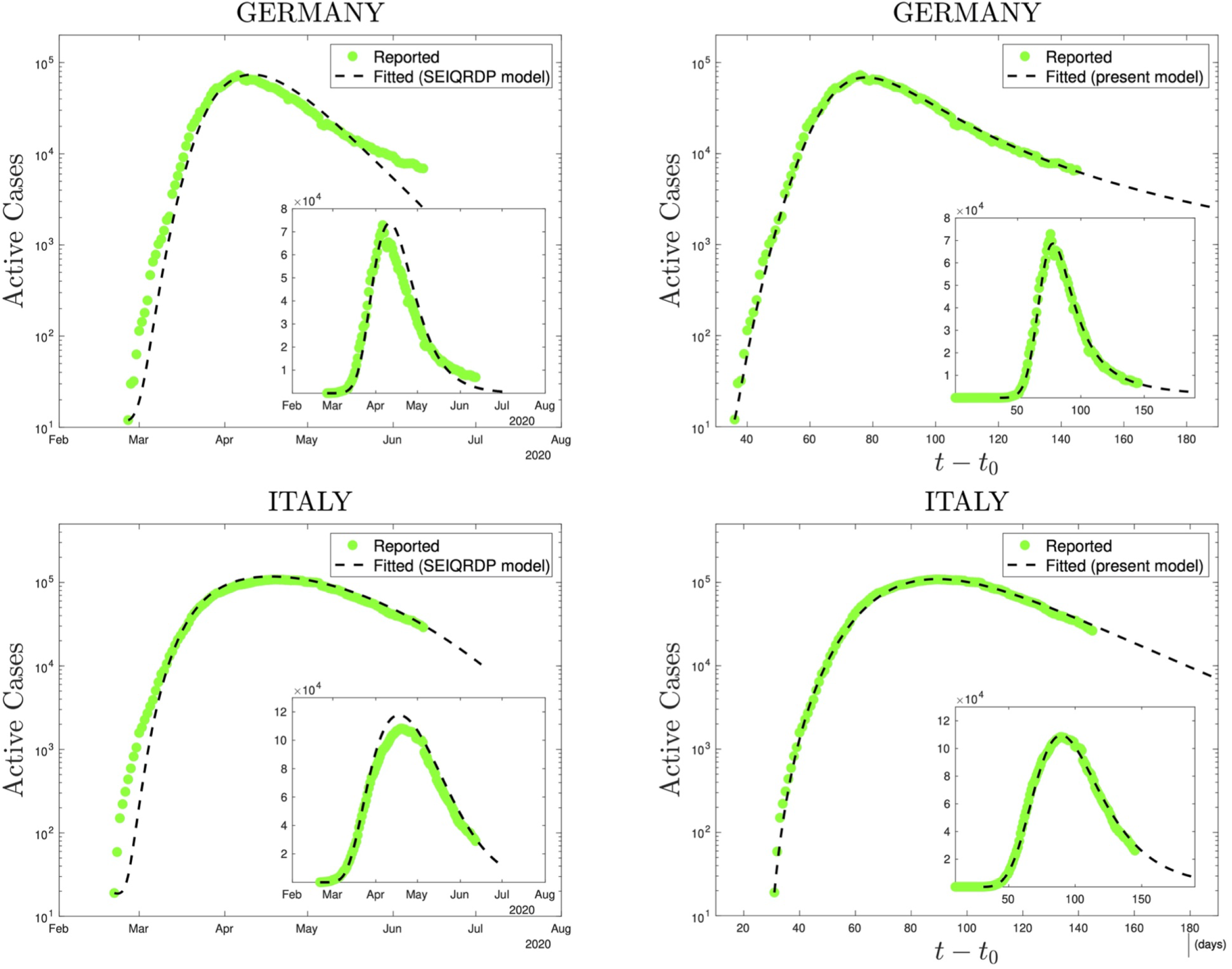
Comparison, using precisely the same reported data (green dots), of the SEIQRDP model (left plots) and the *q*-SEIR model (right plots) for the time series of Germany (from February 26^*th*^ to June 13^*th*^ 2020) and of Italy (from February 21^*st*^ to June 13^*th*^ 2020). To obtain the SEIQRDP fittings (seven linear/bilinear equations satisfying *S* + *E* + *I* + *Q* + *R* + *D* + *P* = *N* and including several fixed as well as two time-dependent coefficients) we have used the online program [47]. To obtain the *q*-SEIR fittings (four not necessarily linear/bilinear equations satisfying *S* + *E* + *I* + *R* = *N*_*eff*_ and including only fixed parameters, two of them being the nonlinear exponents *q*_*up*_ and *q*_*down*_) we have used a standard Least Squares Method (linear scales).

At this stage, let us emphasize a rather interesting fact. Neither the SIR nor the SEIR models distinguish the *dead* from the *recovered*, within the *removed* (R) subpopulation. However, the same values for (*q*_*up*_, *q*_*down*_) fit satisfactorily *both* the numbers of active cases and of deaths per day for a given country, as shown in Figs. 3 and 4. At this point it would be worth noting that although the present model, unlike for example the SEIRD model [48, 49], does not distinguish deaths from healings, the deceased cases will still be roughly proportional to infected people. It is this proportionality, we believe, which makes us to obtain reasonable fits using the variable *I* of the model, again incorporating the proportionality into *N*_*eff*_, hence into *ρ*. It is also evident from these figures that the data for deaths per day are more scattered than those for active cases. This is due to the fact that the real data for the former are much less than for the latter. This is also the reason for the significantly small *ρ* values of deaths per day compared to those of active cases.

## CONCLUSIONS

To conclude let us remind that the *q*-SEIR model recovers, as particular instances, the *q*-SIR model introduced here, as well as the traditional SEIR and SIR ones. It has, however, an important mathematical difference with the usual epidemiological models. Virtually all these models (SIR, SEIR, SAIR, SEAIR, SIRASD, SEAUCR, SEIQRDP) are defined through equations that are multi-linear in their variables, i.e., that are linear in each one of its variables. This multi-linearity disappears in models such as the present *q*-SIR and *q*-SEIR ones if either *q*_*up*_ or *q*_*down*_ differ from unity. Consequently *N*_*eff*_ definitively plays a different role since it sensibly enters within the set of fitting parameters of the *q*-generalized models; its precise interpretation remains to be elucidated, perhaps in terms of the socio-geographical circumstances of that particular region. Last but not least, let us stress that the aim of the present *q*-SEIR model is to *mesoscopically describe a single epidemiological peak*, including its realistic *power-law* growth and relaxation in the time evolution of the number of active cases, and by no means to qualitatively address possibilities such as the emergence of two or more peaks, a task which is (sort of naturally, but possibly less justified on fundamental grounds) attainable within approaches using traditional (multilinear) models where one or more coefficients are allowed to phenomenologically depend on time by realistically adjusting their evolution along the actual epidemics. Alternatively, it is always possible to approach the two-peak case by proposing a linear combination of two *q*-SEIR curves starting each of them at two different values of the departing time (*t*_0_).

## Data Availability

Covid-19 data is available here:
https://data.humdata.org/dataset/novel-coronavirus-2019-ncov-cases

## Acknowledgments

We have greatly benefitted from very fruitful discussions with R. Dickman, T. Pereira and D. Eroglu, as well as from partial financial support by CNPq and Faperj (Brazilian agencies).

